# Highly accurate and precise automated cup-to-disc ratio quantification for glaucoma screening

**DOI:** 10.1101/2024.01.10.24301093

**Authors:** Abadh K Chaurasia, Connor J Greatbatch, Xikun Han, Puya Gharahkhani, David A Mackey, Stuart MacGregor, Jamie E Craig, Alex W Hewitt

**Affiliations:** Menzies Institute for Medical Research, University of Tasmania, Australia; QIMR Berghofer Medical Research Institute, Brisbane, Australia; School of Medicine, University of Queensland, Brisbane, Australia; Faculty of Health, School of Biomedical Sciences, Queensland University of Technology, Brisbane, Queensland, Australia; Lions Eye Institute, Centre for Vision Sciences, University of Western Australia, Australia; Department of Ophthalmology, Flinders University, Flinders Medical Centre, Bedford Park, Australia; Centre for Eye Research Australia, University of Melbourne, Australia

**Author notes:** Correspondence: Abadh K Chaurasia Menzies Institute for Medical Research, University of Tasmania, Australia.

**Keywords:** Artificial Intelligence, Deep Learning, Computer Vision, Glaucoma, Fundus Image, UK Biobank

## Abstract

**Objective:** An enlarged cup-to-disc ratio (CDR) is a hallmark of glaucomatous optic neuropathy. Manual assessment of CDR may be inaccurate and time-consuming. Herein we sought to develop and validate a deep-learning-based algorithm to automatically determine CDR from fundus images.

**Design:** Algorithm development for estimating CDR using fundus data from a population-based observational study.

**Participants:** A total of 184,580 fundus images from the UK Biobank, Drishti_GS, and EyePACS.

**Main Outcome Measures:** The area under the receiver operating characteristic curve (AUROC) and coefficient of determination (R^2^).

**Methods:** FastAI and PyTorch libraries were used to train a convolutional neural network-based model on fundus images from the UK Biobank. Models were constructed to determine image gradability (classification analysis) as well as to estimate CDR (regression analysis). The best-performing model was then validated for use in glaucoma screening using a multiethnic dataset from EyePACS and Drishti_GS.

**Results:** Our gradability model vgg19_bn achieved an accuracy of 97.13% on a validation set of 16,045 images, with 99.26% precision and AUROC of 96.56%. Using regression analysis, our best-performing model (trained on the vgg19_bn architecture) attained an R^2^ of 0.8561 (95% CI: 0.8560-0.8562), while the mean squared error was 0.4714 (95% CI: 0.4712-0.4716) and mean absolute error was 0.5379 (95% CI: 0.5378-0.5380) on a validation set of 12,183 images for determining CDR (0-9.5 scale with a 0.5 interval). The regression point was converted into classification metrics using a tolerance of 2 for 20 classes; the classification metrics achieved an accuracy of 99.35%. The EyePACS dataset (98172 healthy, 3270 glaucoma) was then used to externally validate the model for glaucoma diagnosis, with an accuracy, sensitivity and specificity of 82.49%, 72.02% and 82.83%, respectively.

**Conclusions:** Our models were precise in determining image gradability and estimating CDR in a time-efficient manner. Although our AI-derived CDR estimates achieve high accuracy, the CDR threshold for glaucoma screening will vary depending on other clinical parameters.

**Precis:** Deep-learning-based models can accurately diagnose and monitor glaucoma progression through automated CDR assessment. However, the CDR threshold for glaucoma screening may vary depending on other clinical parameters.

## INTRODUCTION

Primary open-angle glaucoma (POAG) is one of the most common glaucoma subtypes, with a global prevalence of 2.4%.^1^ Accurate detection of glaucoma is essential to prevent irreversible damage to the optic nerve head (ONH), and this often involves clinically assessing the cup-to-disc ratio (CDR). The CDR is a morphological characteristic of the ONH that can estimate the risk of developing glaucoma.^2^ A larger CDR or inter-ocular asymmetry > 0.2 is one of the key risk factors for the development and progression of glaucoma.^3,4,5^ In patients with advanced glaucoma, even small alterations in CDR may lead to considerable loss of retinal ganglion cells.^6^ Furthermore, Genome-wide Association Studies (GWAS) have identified many genetic variants associated with CDR, with a subset of these variants also associated with POAG risk.^7,8,9,10^ Therefore, accurate assessment of CDR is important for glaucoma screening and tracking progression in a clinical setting.

Fundus photography is a non-invasive imaging technique frequently used in glaucoma practice for documenting the condition of the retina, including CDR, neuroretinal rim, disc haemorrhages and longitudinal monitoring of the optic nerve and surrounding retinal structures. Manual assessment of the CDR is also possible but may be challenging and time-consuming for clinicians. Even among glaucoma specialists, CDR measurement is subject to inter-and intra-observer variability.^11,12,13^ Advanced imaging technologies, such as optical coherence tomography (OCT) and confocal scanning laser ophthalmoscopy (CSLO), have been observed to provide different CDR values for an individual.^14,15^ To overcome these difficulties, CDR estimation can be automated through leveraging advances in artificial intelligence (AI) and computer vision.

Several studies have precisely segmented the optic disc and cup with deep-learning-based techniques for calculating CDR using shape-based methods (circular or elliptic ONH) or appearance-based approaches (texture, colour, and intensity of the optic disc).^16,17,18^ Accurately and precisely segmenting the optic cup and disc is challenging due to the variability in ONH morphology, which can be influenced by the patient’s ethnicity, age, disease conditions and image quality.^19,20,21^ Training a segmentation model requires a large amount of data for the optic cup and disc boundaries manually labelled by experts.^22^ A limited number of studies have used convolutional neural network (CNN)-based regression analysis to estimate CDR using fundus image data.^23,24,25^ Therefore, we developed and validated a CNN-based regression model on a large cohort of retinal images to compute CDR without applying the segmentation techniques for diagnosing glaucoma in clinical and community settings.

## METHODS

### Study Design

A comprehensive overview of this study is shown in **Figure 1**. An extensive dataset from the UK Biobank (UKBB) was used, with each retinal image in the study independently assessed and graded by two ophthalmologists (AWH and JEC). This research utilised the CNN-based models for classification and regression analyses to determine the gradability and estimate the CDR from 80,225 and 68,695 coloured fundus images, respectively.

**Figure 1:**
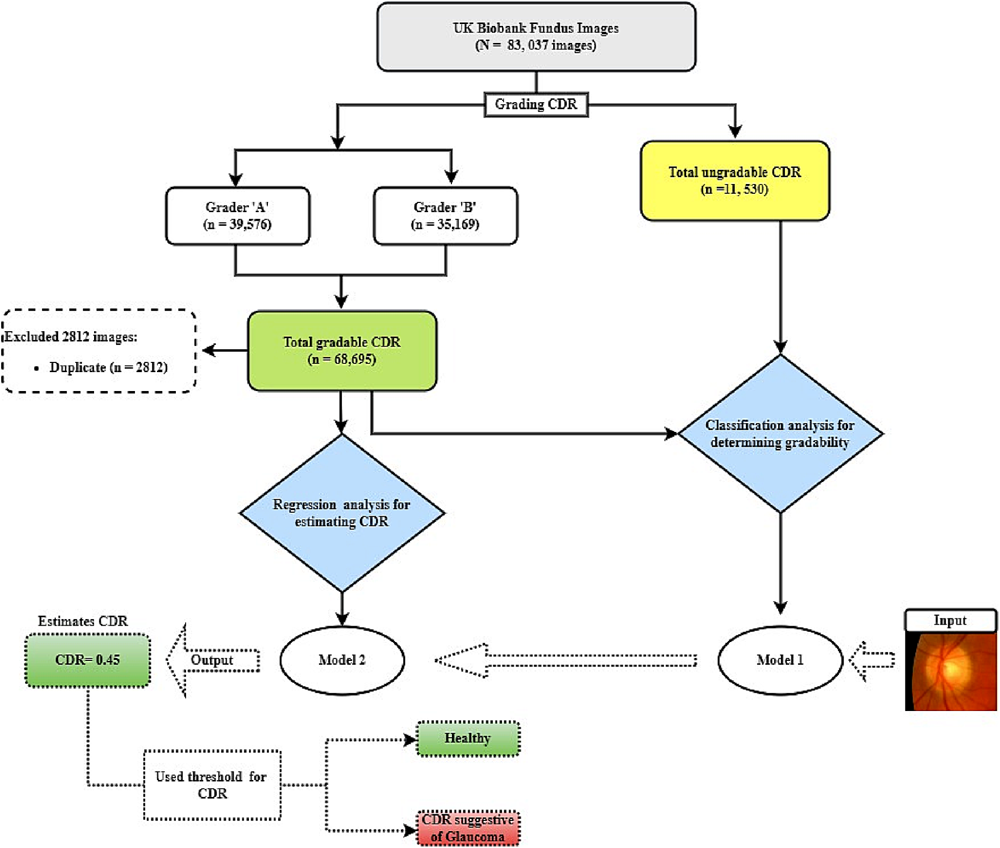
The study flow diagram for diagnosing glaucoma using classification model 1 for gradability assessment and regression model 2 for estimating the CDR on UK Biobank retinal photographs.

The UKBB study received ethical approval from the local research ethics boards, and all participants gave informed written consent. This study’s procedures were performed in accordance with the ethical guidelines for medical research as described by the World Medical Association Declaration of Helsinki.

### Participants and Imaging Modality

#### Training Dataset

All the participants’ data were from the UKBB dataset, which is a large, population-based observational study in the United Kingdom that began in 2006 and recruited over 500,000 participants aged between 40 to 69 years at the time of recruitment—67,040 participants had at least one gradable retinal image.^26,27^ The images were available from both left and right eyes from baseline and repeat assessment visits. The retinal images were obtained with a Topcon 3D OCT 1000 Mark II (Topcon Corp., Tokyo, Japan).^28^ This study included both eyes of the participants during two visits, irrespective of ancestry.

#### External Validation

The models were externally validated on the EyePACS dataset—an excellent and reliable resource for creating AI-powered tools to diagnose glaucoma.^29^ This extensive collection of 101,442 fundus images represents a diverse population of 60,357 individuals who visited various centres across the EyePACS network in the United States.^30^ This dataset is of exceptional quality and can be relied upon to deliver accurate results. Our models were also validated on another geographic and ethnic dataset of Drishti (India)^31^ to see how models are consistent and effective across different populations. It includes 70 images from people diagnosed with glaucoma and 31 normal images, all with a resolution of 2047×1760 pixels in PNG format.

#### Image Grading and Pre-processing

In our previous study, two fellowship-trained ophthalmologists independently viewed and graded the retinal images.^27^ All the fundus images were cropped to a pixel ratio of 1080 x800 before the training or validation of the gradability assessment. However, for regression analysis, we further cropped and removed the non-informative areas, background or margin around the main content of the images based on their pixel intensity using the OpenCV library.^32^ The final downsized images were 512 x 512 pixels. As undertaken previously, retinal photographs with low-quality images (ungradable and artefacts) were removed.^33,34^

#### Model Selection

Twelve pre-trained models from the FastAI (TorchVision)^35,36^ were utilised and tested on grader “A” data (larger dataset) for CDR estimation using regression analysis. The potential model (vgg19) with batch normalisation (bn) layers was selected based on the model’s performance (**Supplementary** Figure 1) for both classification and regression analyses. The vgg19_bn pre-trained model was initially trained on ImageNet.^37^ This pre-trained model automatically learns high-level features from a large variety of images; thus, it offers a robust feature extractor for classification and regression tasks. We trained and fine-tuned the CNN regression model on a combined dataset of 68,695 images to estimate CDR. The models for detecting glaucoma, using the overall cut-off CDR (≥0.60),^38^ were extensively validated on two publicly available datasets including different ethnicities.

### Deep-Learning Algorithm and Training

All the models were trained via the FastAI Framework—a deep-learning library built on PyTorch that provides a high-level application programming interface (API) and allows deep-learning architectures to be trained quickly to achieve state-of-art results.^39^ We used two separate CNN-based models from the FastAI library for the gradability assessment of CDR and regression analysis for estimating CDR using fundus images. The binary cross entropy was utilised for the loss function for the gradability task, and the MSE loss function was used for regression analysis.

Image gradability was defined as binary classification (gradable or ungradable) based on the CDR that clinicians can obtain. The fundus images were randomly (80:20) separated into two groups, to train and validate the models for classification and regression analyses. The classification model was trained on a dataset of 64,180 fundus images, while the regression model was trained on a dataset of 48,734. The validation sets for the classification and regression model comprised 16,045 and 12,183 images, respectively.

The class weights were balanced for the gradability task, and in-built data augmentation techniques were implemented for both classification (softmax probability exceeding 0.5 was designated as gradable) and regression analysis to improve the accuracy for generalising the models’ utility.^40^ The augmentation parameters are illustrated in **Supplementary Table 1**. The training was completed in two steps: in the first step, we used fine-tuning with frozen layers of the pre-trained weights (transfer learning) for 5-10 epochs using Valley to find the learning rate—validation loss was monitored, and the task was executed until it reached the point where validation loss stopped decreasing. In the next step, the model unfroze all their layers and used a 1-cycle policy for ten epochs with the ‘slice’ function to implement a strategy known as discriminative learning rates during the model training.^39,41^ The callback function was employed to monitor the validation loss during the training and stop training if the loss failed to improve by at least 0.1 (minimum delta) at patience 2 for a maximum of 10 epochs. The regularisation technique (weight decay= 1e-3) was also operated to prevent overfitting.

The experiment was conducted on virtual Ubuntu (22.04) desktop with NVIDIA A100 with 40GB of GPU RAM at Nectar Research Cloud^42^ using Python (3.10.6) programming language with PyTorch (2.0.0+cu117), FastAI (2.7.12), TorchVision (0.15.1+cu117), Matplotlib (3.5.1), and Scikit-learn (1.2.2) libraries.^43,44,45^

### Model Evaluation

The models were evaluated on the area under the receiver operating characteristic curve (AUROC), sensitivity, specificity, precision, recall, and F1-score for gradability assessment, and mean absolute error (MAE), mean squared error (MSE), correlation Coefficient (R), and coefficient of determination (R^2^) were used for regression analysis with a 95% confidence interval (CI) for each outcome.^46^ These metrics assessed model performance by quantifying the difference between actual (graded by clinicians) and predicted CDR by the models. Furthermore, the classification metrics were measured by setting a tolerance bin around each regression point and seeing if the truth value fell within tolerance.

## RESULTS

### Training Dataset

As described previously, a total of 83,037 fundus images from the UKBB fundus dataset were evaluated and had CDR assessed.^27^ Of these images, 68,695 (82.73%) were gradable, and 2,812 (3.38%) duplicate images were removed. Two independent graders exhibited a strong positive correlation (Pearson correlation coefficient = 0.70) and high consistency (Intraclass Correlation Coefficient, ICC3k = 0.82) on a shared set of 2,812 images (**Supplementary** Figure 2).

### Gradability Assessment

Our classification model obtained a high accuracy of 97.13% on the internal validation set of 20% (16,045) of the total data (80,225) for gradability assessment: gradable and ungradable. The CI with 95% for each metric were estimated through Bootstrap resampling^47^ with 4000 iterations to ensure the reliability of the results (**Table 1**). The model underwent testing on random images from the validation set, and the outcome from the classification model is visualised in **Supplementary** Figure 3.

**Table 1:** Gradability performance of classification model (model 1) for both gradable and ungradable fundus images based on CDR.

| Classification Metrics | Gradable<br>(n =13,717)<br>Mean [95% CI] | Ungradable<br>(n =2328)<br>Mean [95% CI] |
| --- | --- | --- |
| **AUROC** | 0.9656 [0.9614, 0.9697] | 0.9656 [0.9613, 0.9699] |
| **Accuracy** | 0.9713 [0.9688, 0.9738] | 0.9712 [0.9687, 0.9738] |
| **Sensitivity (Recall)** | 0.9736 [0.9710, 0.9763] | 0.9575 [0.9499, 0.9652] |
| **Specificity** | 0.9574 [0.9491, 0.9657] | 0.9736 [0.9709, 0.9763] |
| **Precision** | 0.9926 [0.9912, 0.9941] | 0.8605 [0.8471, 0.8738] |
| **F1-score** | 0.9830 [0.9815, 0.9846] | 0.9063 [0.8981, 0.9145] |

n = Number of fundus images, CI = Confidence Intervals

### Regression Analysis

A total of 68695 images were retrieved for regression analysis. Of these, 80% (54956) were used for training the vgg19_bn model, while the remaining 20% (13739) were reserved for model validation. We excluded 2812 duplicate images before training the model, which achieved an R^2^ of 0.71 on the validation set. To further enhance its performance, we eliminated 7780 images from the top loss predicted by the model using the ImageClassifierCleaner() from FastAI library.^48^ We then re-trained the regression model on the final dataset of 60917 images, with 80% used for training and 20% for validation, resulting in a 15% improvement in the R^2^. Our regression metrics are illustrated in **Table 2**. The Bland-Altman plot displays the accuracy of the CDR estimation on a validation dataset. The plot includes the corresponding CDR ground truth, which ranges from 0 to 9.5 with 0.5 intervals. Figure 2 **(a)** exhibits the mean offsets, agreement limits with a 95% CI.

**Figure 2:**
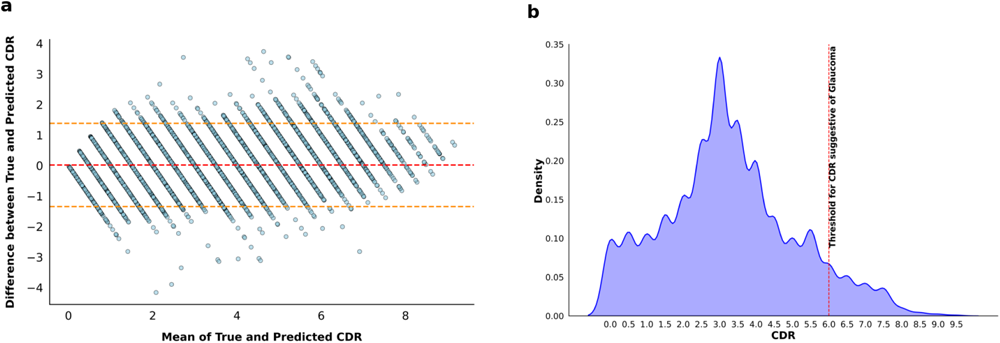
Comparative analysis and final dataset distribution. The Bland-Altman plot displays the CDR estimation on the validation dataset with the corresponding CDR ground truth shown in (**a**). The mean difference between true and predicted CDR was 0.022 (95% CI: 0.009-0.034). The red dashed line represents the mean difference, while the yellow dashed lines represent the 95% CI for the mean difference for upper and lower limits. The distribution of the CDR in the final analysis data is graphically presented in (**b**).

**Table 2:** Regression metrics for estimating CDR.

| Regression Metrics | Value |
| --- | --- |
| **MSE** | 0.4714, 95.0% CI: [0.4712, 0.4716] |
| **MAE** | 0.5379, 95.0% CI: [0.5378, 0.5380] |
| **R<sup>2</sup>** | 0.8561, 95.0% CI: [0.8560, 0.8562] |
| **R** | 0.9258, 95.0% CI: [0.9258, 0.9259] |

**MSE** = Mean Squared Error, **MAE** = Mean Absolute Error, **R<sup>2</sup>** = Coefficient of Determination **R** = Correlation Coefficient, **CI** = Confidence Intervals

The mean CDR values from graders A and B were 3.63 and 3.06, respectively, but the final mean and standard deviation for the analysed data were 3.26 and 1.8. Figure 2 **(b)** visualises the distribution of CDR from the final analysed dataset, while **Supplementary** Figure 4 shows the random images taken from the validation set to estimate CDR by the regression model.

The classification metrics were also assessed using a tolerance-based approach, wherein a predefined tolerance bin was established around each regression point. Subsequently, the true value was evaluated to determine if it fell within the established tolerance bin. Thus, we examined the model’s classification metrics using four tolerance thresholds (0.5, 1.0, 1.5, and 2.0) around the ground truth values (**Supplementary Table 2)**. The regression point was converted into classification metrics with a tolerance of 2 for 20 classes, and the classification metrics achieved an accuracy of 99.35%. This approach facilitated the rigorous evaluation of the model’s accuracy in correctly assigning class labels; these twenty classes are displayed in the confusion matrix in Figure 3 **(a)**.

**Figure 3:**
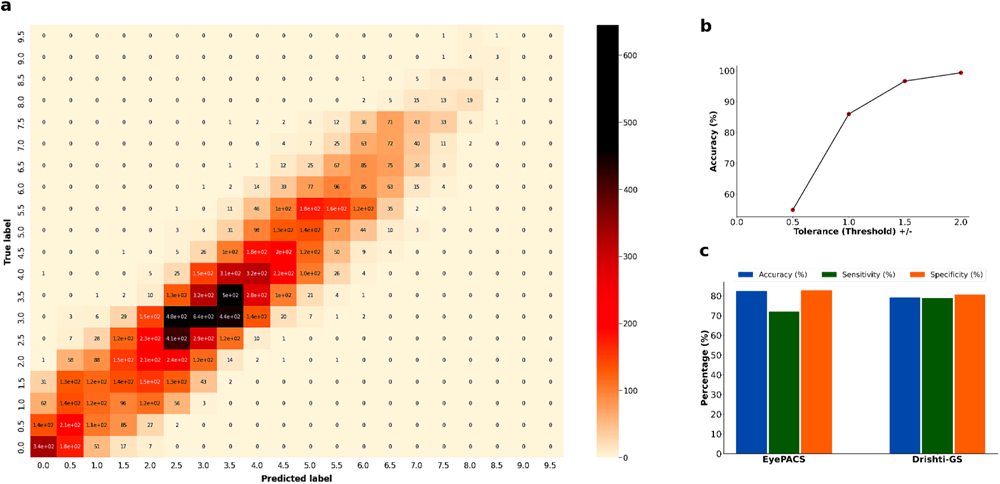
The confusion matrix for CDR classification is illustrated in (**a**). In contrast, (**b**) presents the conversion of regression points into classification metrics utilising a particular threshold. Publicly available datasets were used for external validation of the glaucoma screening at the globally accepted cut-off threshold, as shown in (**c**).

### Glaucoma Screening

Our models were externally validated on EyePACS and Drishti datasets with promising results for screening glaucoma. On EyePACS, our models achieved 82.49% accuracy, 72.02% sensitivity, and 82.83% specificity. On Drishti, the models achieved 79.21% accuracy, 78.87% sensitivity, and 80.65% specificity. These models predicted less than 1% and 0% ungradable images on the EyePACS and Drishti datasets, respectively, as shown in **Supplementary Table 3**.

Although we extensively analysed the EyePACS and Drishti datasets, the optimal CDR threshold for detecting glaucoma was 0.56 (**Supplementary** Figures 6 **& 7**) using Receiver Operating Characteristic (ROC) curve analysis.^49^ The AUROC was 0.87, suggesting this model can distinguish fundus images with and without glaucomatous optic discs.

## DISCUSSION

In this study, we conducted the first regression analysis on 184,580 fundus images utilising deep-learning architectures to estimate the CDR without employing segmentation techniques from image gradability to glaucoma screening in a single study. Our classification model was first trained for the gradability task and successfully predicted gradable fundus images based on CDR with remarkable accuracy, precision, and recall. Our results outperformed Yuen et al.’s fundus image quality assessment study, which reported an accuracy of 92.5%, a sensitivity of 92.1%, and a specificity of 98.3% on internal validation with 11.6 times fewer validation data than ours.^50^ However, both studies demonstrate that the deep-learning-based model has the potential to grade fundus images accurately.

Accurate estimation of the CDR is required in diagnosing and monitoring glaucoma. Conventionally, AI-based CDR estimation relies on segmentation techniques.^51,52,21,53,54^ This technique can be computationally expensive and prone to errors in accurately estimating CDR. Aljazaeri et al. reported that a CNN-based regression model performs better than the standard segmentation technique for calculating CDR.^55^ Our study utilised a deep learning-based model to directly estimate CDR from the fundus images using feature extraction by pre-trained weights (vgg19_bn) without applying segmentation techniques. This approach provides a more efficient and reliable method for estimating CDR.

Our regression model performed exceptionally well on a large dataset of retinal images from the UKBB dataset, achieving an R^2^ value of 0.8561, with a 95% confidence that the actual CDR value lies between 0.8560 and 0.8562 on the validation set (12,183). It is worth mentioning that Hemelings et al. also used CNN regression analysis to estimate CDR from fundus images, and their findings were notable. However, our results surpassed theirs with an R^2^ value of 86% compared to their 77% [95% CI 0.77-0.79] on the test set of 4,765 for CDR estimation. We observed that their mean of CDR was 0.67, approximately twice our dataset’s CDR mean.^23^ Their different regression model exhibited a strong Pearson’s correlation coefficient (R) of 0.88 when predicting the CDR against expert grader.^25^ Another study reported that the model achieved an R of 0.89 on a dataset of 2,115 fundus images from the UKBB cohort.^56^ In contrast, our model achieved a higher R of 0.93, indicating a strong correlation between the predicted and expert-graded CDR. This enhanced performance improves our model’s efficiency and precision for estimating CDR, which could be pivotal in glaucoma decision-making in clinical settings.

Using our regression model, we can accurately determine the glaucomatous ONH features (*i.e.* the CDR) from the retinal fundus images, offering a cost-effective and time-efficient alternative to labour-intensive and potentially inconsistent expert labelling required in downstream studies such as GWAS.^8,56^ Our combined models could assess the gradability, CDR estimation, and glaucoma classification within 0.3 seconds per retinal image. Contrastingly, glaucoma screening based on CDR using segmentation techniques takes 11 seconds per image.^57^ We also noticed that our regression model could accurately estimate the CDR even from poor-quality retinal images (**Supplementary** Figure 5). This research will be a game-changer in scenarios where fundus images are blurred or poor quality for glaucoma assessment, especially in cases involving patients with cloudy media.

Our diagnostic models for glaucoma exhibit great potential in a clinical setting. Despite this, the screening of glaucoma on a global scale poses challenges due to the variability of optic disc sizes among different ethnic groups.^58,59^ This creates obstacles in standardising screening techniques using computer vision based on CDR. Hemelings et al. reported an impressive AUROC for external validation but altered in both sensitivity and specificity values across the different datasets when using a fixed threshold (0.7) for glaucoma.^25^ The highest specificity was observed in the REFUGE1 dataset at 0.99 [0.98-0.99], whereas the PAPILA dataset exhibited the lowest specificity at 0.70 [0.63-0.76]. Regarding sensitivity, the GHS, PAPILA, and AIROGS datasets achieved the highest value of 0.94, with the ORIGA dataset presenting the lowest sensitivity at 0.68 [0.61-0.75]. They also validated the model externally on the EyePACS dataset; sensitivity was achieved 0.68 and 0.89, with thresholds set at 0.82 and 0.72, respectively. Bhuiyan et al. noted 80.11% of sensitivity and 84.96% of specificity on the ORIGA dataset for screening glaucoma suspects using retinal images, with a cut-off point of CDR >0.5.^60^ Consequently, depending exclusively on CDR for glaucoma screening may overlook the complexity of glaucoma (**Supplementary Table 3**). A robust and comprehensive screening model for glaucoma at a population level requires integrating glaucomatous phenotypic and genotypic information with advanced AI algorithms.

This work has some important limitations. The dataset used in the training of the models was mainly from the healthy Caucasian population—the mean of CDR was 0.37—potentially biased for glaucomatous discs; the distribution of CDR can be seen in Figure 2 **(b).** The sample we used to train our models may not represent other age groups or populations from different ethnic or geographical backgrounds. Second, we downsized the input images (224 x 224 x 3) to train the regression model, which could lose some meaningful information for accurately estimating the CDR. We dropped out approximately 11% of gradable images from top loss; that could potentially make more imbalance classes.

In summary, we developed a fully automatic end-to-end computer vision model for estimating CDR from the fundus images using CNN regression analysis for glaucoma care. The models were externally validated on two publicly available databases for glaucoma screening based on CDR. Our results demonstrated that the models perform very well for both gradability and estimating CDR in a time-efficient manner—this could be implemented in clinical settings. However, a generalised model for screening glaucoma based on CDR is challenging due to uncertainty in what the optimal CDR cut-off should be for diagnosing glaucoma.

## Data Availability

All data produced in the present study are available upon reasonable request to the authors.

## Acknowledgements

This research has been conducted using the UK Biobank Resource under Application Number 25331. SM, DAM, and AWH acknowledge Program grants from the National Health and Medical Research Council (GNT1150144) and Centre of Research Excellence (1116360) funding from the Australian National Health and Medical Research Council (NHMRC). SM and DAM are supported by research fellowships from the Australian National Health and Medical Research Council (NHMRC). PG (#1173390) and AWH are supported by NHMRC Investigator Grants. Also supported by Research Training Program Scholarship from the University of Tasmania (AKC).

