## Supplementary material for "Highly accurate and precise automated cup-to-disc ratio quantification for glaucoma screening": Supply

**Affiliations:**

***Correspondence:**

Abadh K Chaurasia

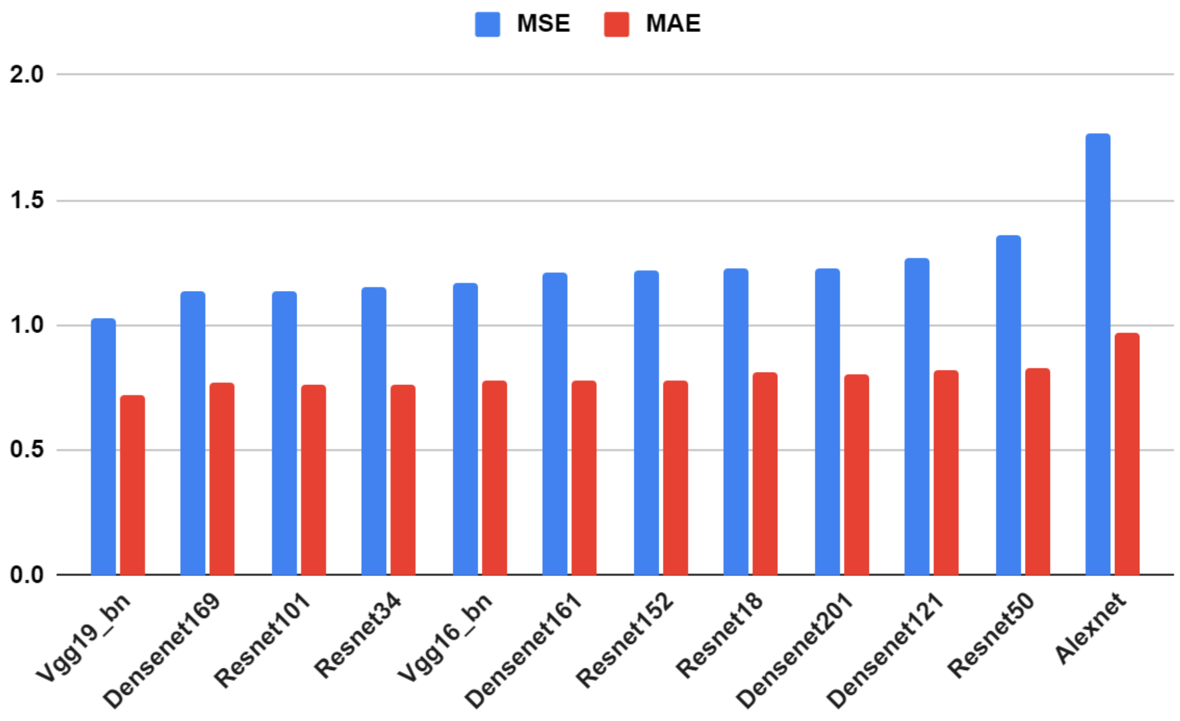


**Supplementary Figure 1:** *All the models were tested on grader "A" data for determining the potential model based on Mean Squared Error (MSE) and Mean Absolute Error (MAE).*

**Supplementary Table 1:** *Summary of the data block and batch transforms parameters for classification and regression tasks.*

**i). Data Block:**

| **Parameter** | **Classification Task** | **Regression Task** |
|---|---|---|
| blocks | (ImageBlock, CategoryBlock) | (ImageBlock, RegressionBlock(n_out=1)) |
| get_items | get_image_files | - |
| splitter | RandomSplitter(valid_pct=0.2) | RandomSplitter(valid_pct=0.2) |
| get_x | - | get_x |
| get_y | lambda x: str(parent_label(x)) | get_y |
| item_tfms | Resize(512) | Resize(512) |

**ii). Batch Transforms**

| **Transformation** | **Classification Task** | **Regression Task** |
|---|---|---|
| aug_transforms | size=224, min_scale=0.75, max_lighting=0.05, do_flip=True, flip_vert=False, max_rotate=15, max_warp=0.0, p_affine=0.8, max_zoom=0.1, p_lighting=0.8 | size=224, min_scale=0.75, max_lighting=0.1, do_flip=True, flip_vert=False, max_rotate=15, max_warp=0.1, p_affine=0.8, max_zoom=0.15, p_lighting=0.8 |
| Resize | 224, 224 | 224, 224 |
| Normalize | Normalize.from_stats(*imagenet_stats) | Normalize.from_stats(*imagenet_stats) |

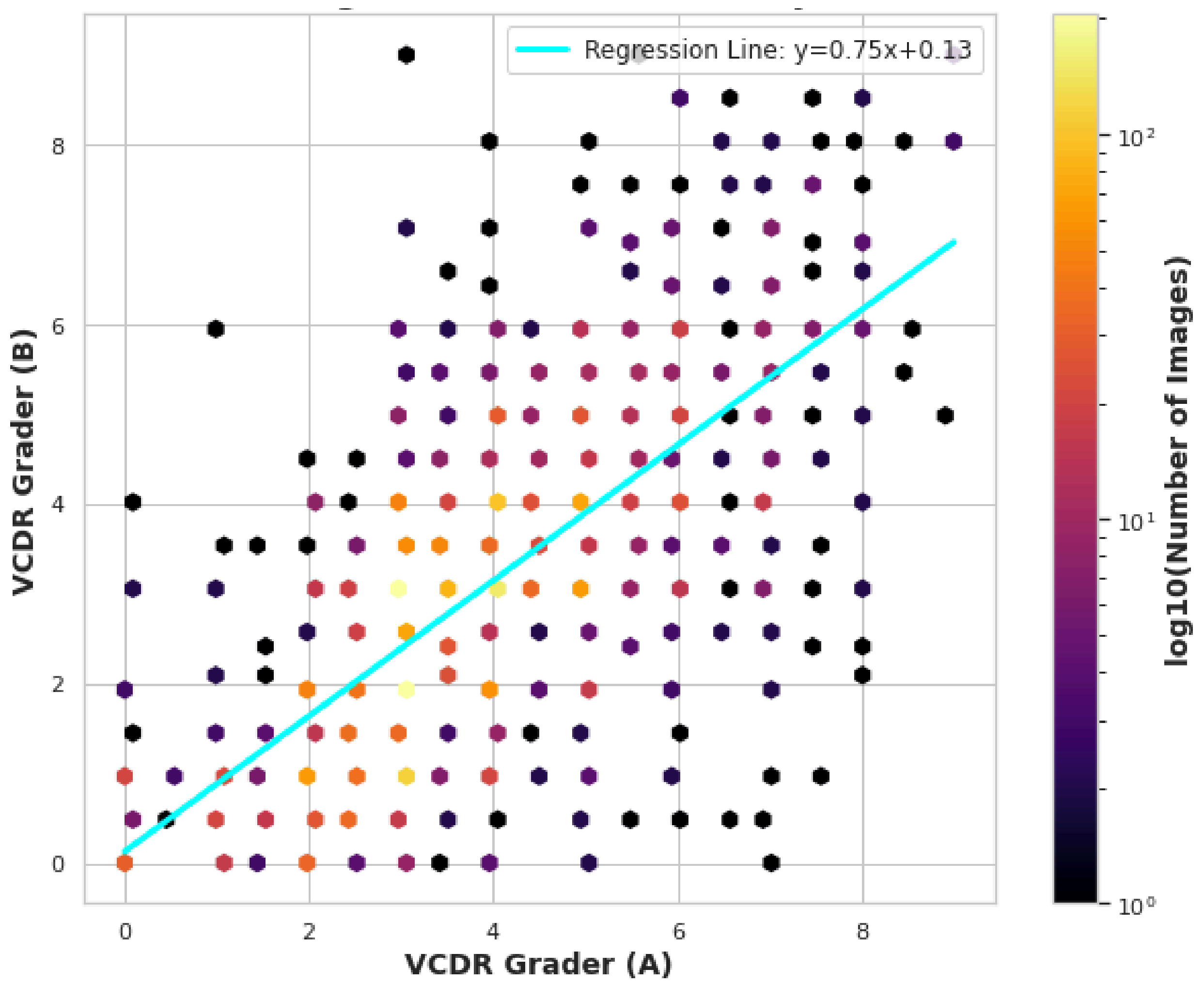


**Supplementary Figure 2:** *Comparison of CDR grading between two graders: Correlation Analysis. The hexbin plot shows the distribution of CDR values determined by graders 'A' and 'B'. The regression line highlights the relationship between the two graders, with an ICC of 0.82 and a Pearson correlation coefficient of 0.70.*

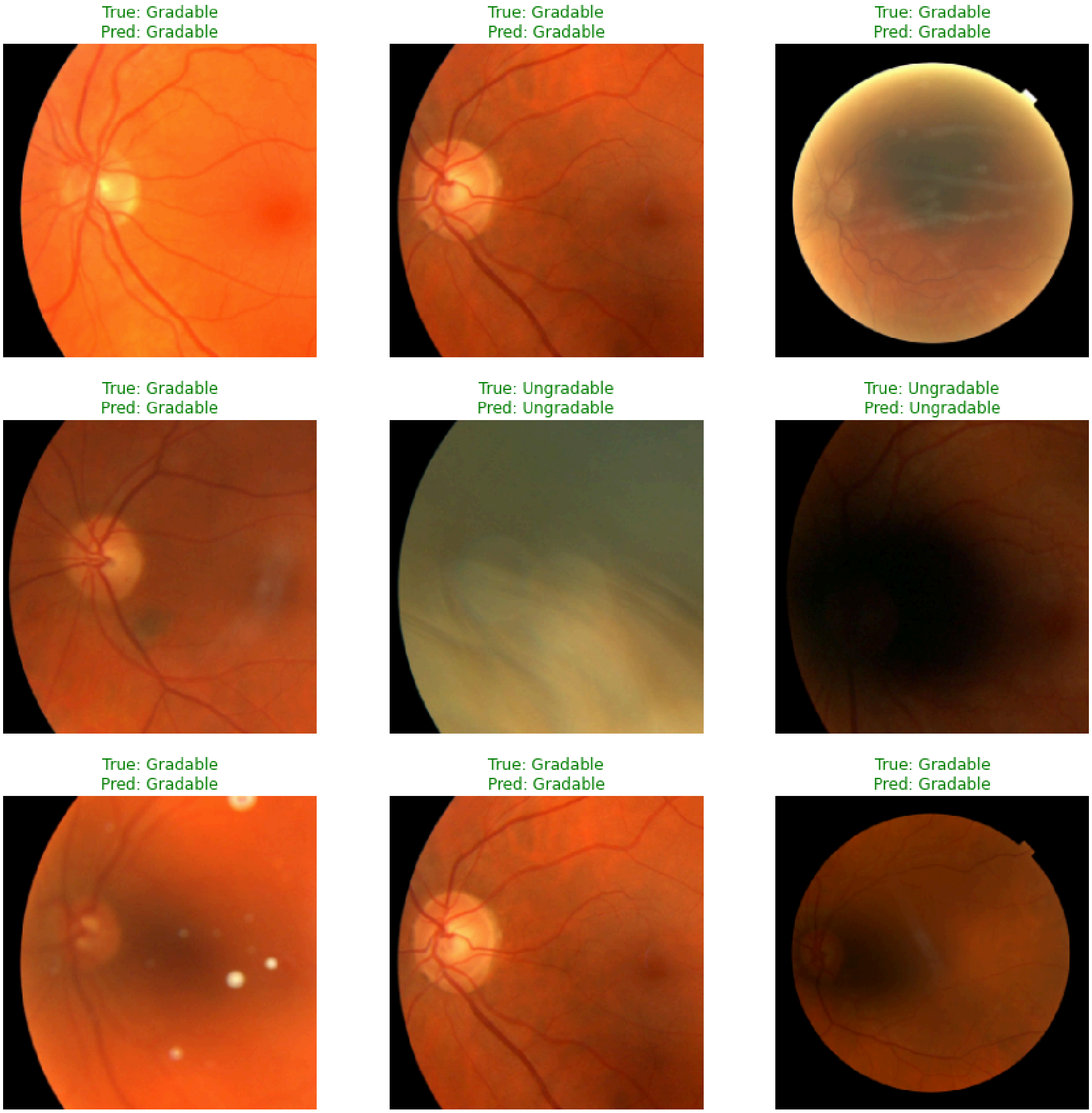


**Supplementary Figure 3:** *Random prediction from the classification model (Model 1), gradable vs ungradable fundus images.*

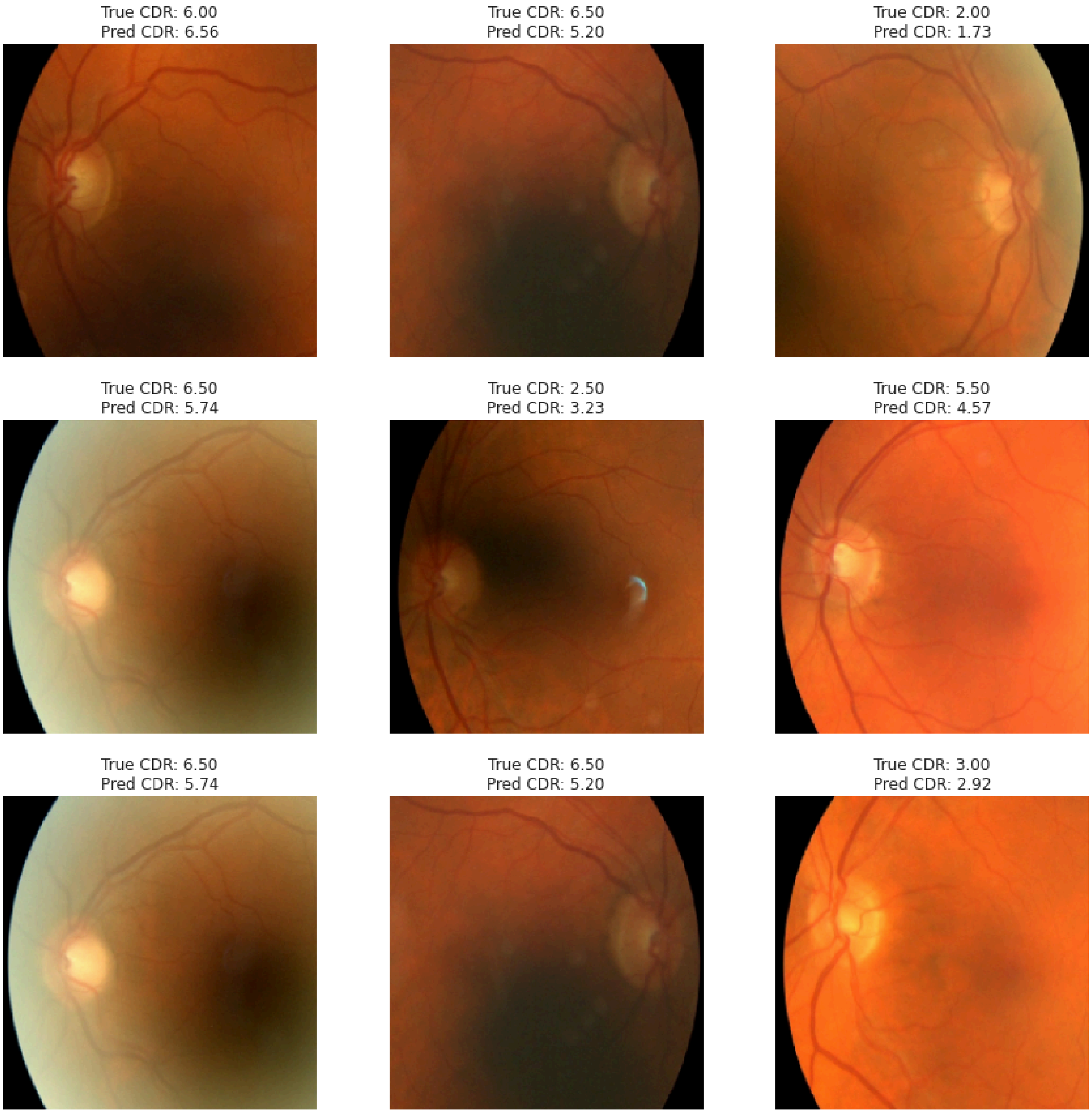


**Supplementary Figure 4:** *Random prediction of CDR from the regression model (vgg19_bn) using fundus images.*

**Supplementary Table 2:** *The conversion of regression points into classification metrics utilising variable thresholds.*

| Threshold Tolerance (+/-) | Accuracy (%) |
|---|---|
| **0.5** | **54.90** |
| **1.0** | **85.98** |
| **1.5** | **97.18** |
| **2.0** | **99.35** |

**Supplementary Table 3:** *External validation on publicly available datasets for glaucoma screening at various cut-off thresholds for glaucoma.*

| Dataset | Number of Images (Healthy=H, Glaucoma=G) | Cut-off Threshold | Performance Metrics (%) | | | Predicted ungradable Images | Time Taken (seconds) |
|---|---|---|---|---|---|---|---|
| | | | Accuracy | Sensitivity | Specificity | | |
| **EyePACS** | H=98,172, G=3270 | 0.5 | 61.05 | 93.28 | 60.00 | 955 | 13251.07 |
| **Drishti-GS** | H=31, G=70 | 0.5 | 84.16 | 90.00 | 70.97 | 0 | 35.63 |
| **EyePACS** | H=98,172, G=3270 | 0.6 | 82.49 | 72.02 | 82.83 | 955 | 26789.02 |
| **Drishti-GS** | H=31, G=70 | 0.6 | 79.21 | 78.87 | 80.65 | 0 | 47.37 |
| **EyePACS** | H=98,172, G=3270 | 0.7 | 94.92 | 30.64 | 97.01 | 955 | 27177.49 |
| **Drishti-GS** | H=31, G=70 | 0.7 | 64.36 | 50.00 | 96.77 | 0 | 35.46 |

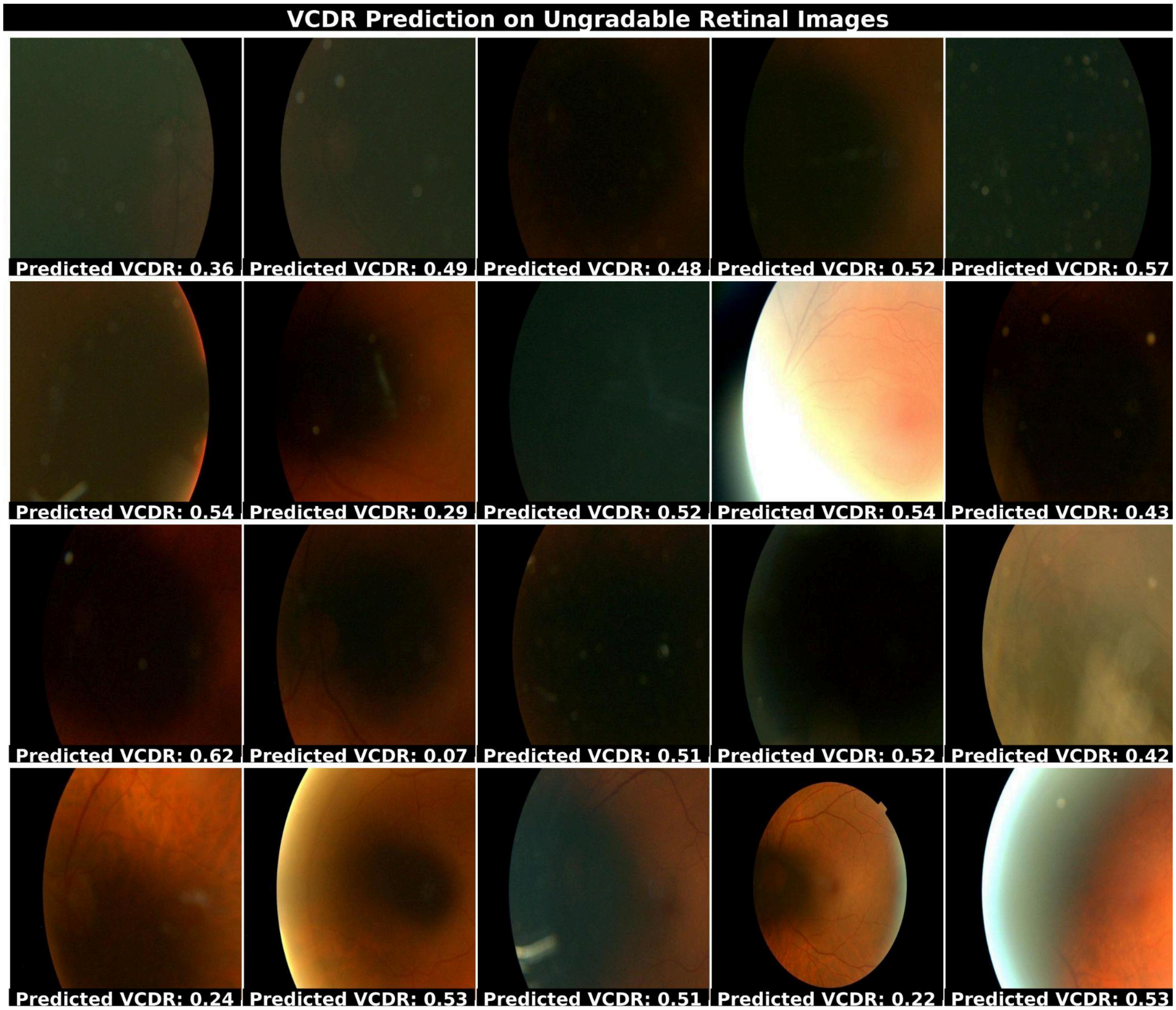


**Supplementary Figure 5:** *VCDR prediction on ungradable fundus images.*

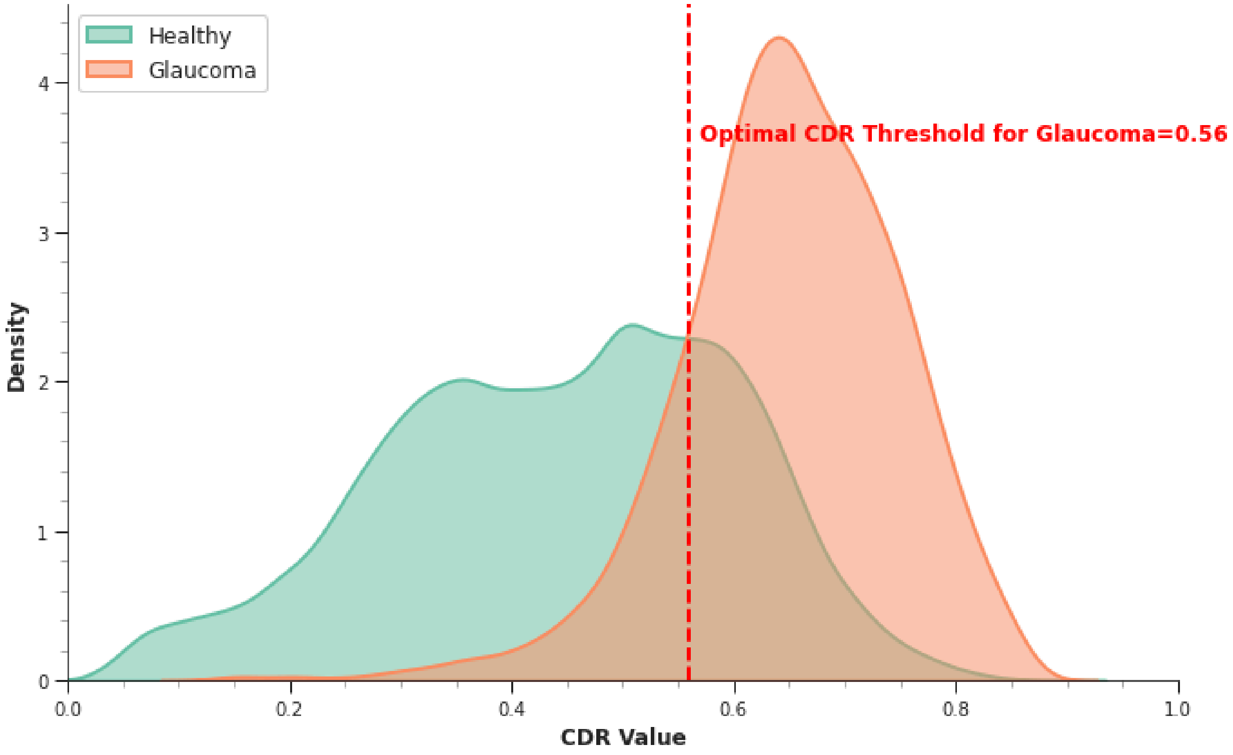


**Supplementary Figure 6:** *Comparative distribution of CDR for glaucoma and healthy fundus images with optimal CDR threshold for diagnosing glaucoma.*

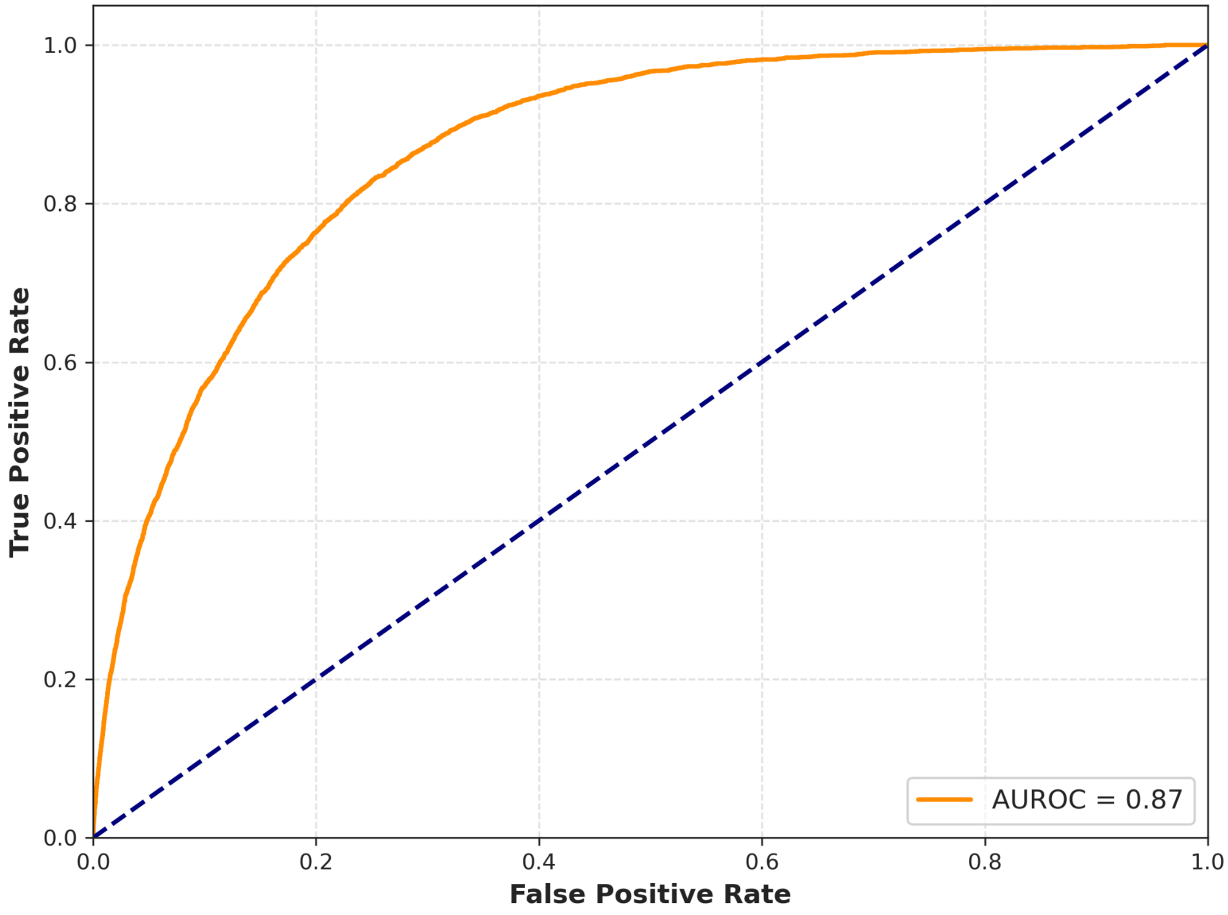


**Supplementary Figure 7:** *Receiver operating characteristic (ROC) curve analysis for determining optimal CDR threshold for diagnosing glaucoma.*
